# Emergence of the D614A mutation in the spike protein of SARS-CoV-2: Imported cases to the South Korea

**DOI:** 10.1101/2020.09.04.20184721

**Authors:** Ae Kyung Park, Jin Sun No, Eunkyung Shin, Yoon-Seok Chung, Il-Hwan Kim, Byung-Hak Kang, Gi-Eun Rhie, Heui Man Kim, Jeong-Min Kim, Ye Eun Park, Min Joon Kim, Jae Sun Park, Cheon-Kwon Yoo, Junyoung Kim

## Abstract

We identified imported cases of a novel D614A mutation in the spike glycoprotein of SARS-CoV-2 in the South Korea. SARS-CoV-2 harboring the novel mutation was isolated from returning travelers from Uzbekistan. These results emphasized the possibility that new mutations have emerged in that area.

---

Since a new novel beta-coronavirus, SARS-CoV-2, was first reported in December 2019, there has been ongoing explosive spread of this virus throughout the world, severely threatening human health (1). South Korea was one of the first countries to report a significant Coronavirus Disease-19 (COVID-19) outbreak; however, the epidemic curve has been quickly flattened due to the outstanding response of the Korean government (2). Currently, community spread of Covid-19 has slowed in the South Korea. On the other hand, more than half of confirmed cases were related to travel (2). The Korea Centers for Disease Control and Prevention (KCDC) has begun to conduct whole-genome sequencing (WGS) of COVID-19 isolated from travelers to obtain real-time genomic epidemiology data, rapidly monitor how the virus mutates and investigate sequence variations.

On July, 2020, a man who entered Korea from Uzbekistan (Uzbekistan nationality) and had symptoms of cough, fever, and headache was diagnosed with COVID-19 (3). For WGS, total RNA was extracted, and cDNA was amplified using primer pools (https://artic.network/ncov-2019). Libraries were prepared using the Nextera DNA Flex Library Prep Kit (Illumina, USA), and sequencing was performed on a MiSeq instrument with 2 x 75 base pairs using a MiSeq reagent kit V3 (Illumina, USA). The reads were mapped to the reference genome MN908947.3 using CLC Genomics Workbench version 20.0.3 (CLC Bio, Denmark). A total of 663,666 (98.50%) out of a total of 673,800 reads mapped to the reference genome. We obtained 99.72% genome coverage at the median genome coverage depth of 1554× (Supplementary Table 1). The genomic sequences of the SARS-CoV-2 strain were deposited in GISAID (Accession no. EPI_ISL_516783, EPI_ISL_516784, EPI_ISL_516785). Using the reference genome, single-nucleotide variants (SNVs) were called using the BioNumerics version 7.6 SARS-CoV-2 plugin (Applied Maths, Belgium).

We identified 14 polymorphic sites in the genome, which corresponded to all SNVs (Table 1). In total, 13 appeared within the gene-coding regions. Of note, we identified a GAT→GCT (A23403C) nucleotide change, which leads to a nonsynonymous change (D614A) in the spike (S) glycoprotein. Based on 77,577 viral genomes downloaded on August 5, 2020, from the GISAID database (4), the DNA sequences corresponding to the S protein of each genome were extracted and aligned to the mutated S protein (D614A) (Table 1). The alignment results indicated that the D614A mutation of the S protein identified in this study was a novel mutation that has not been identified previously.

**Table 1.**
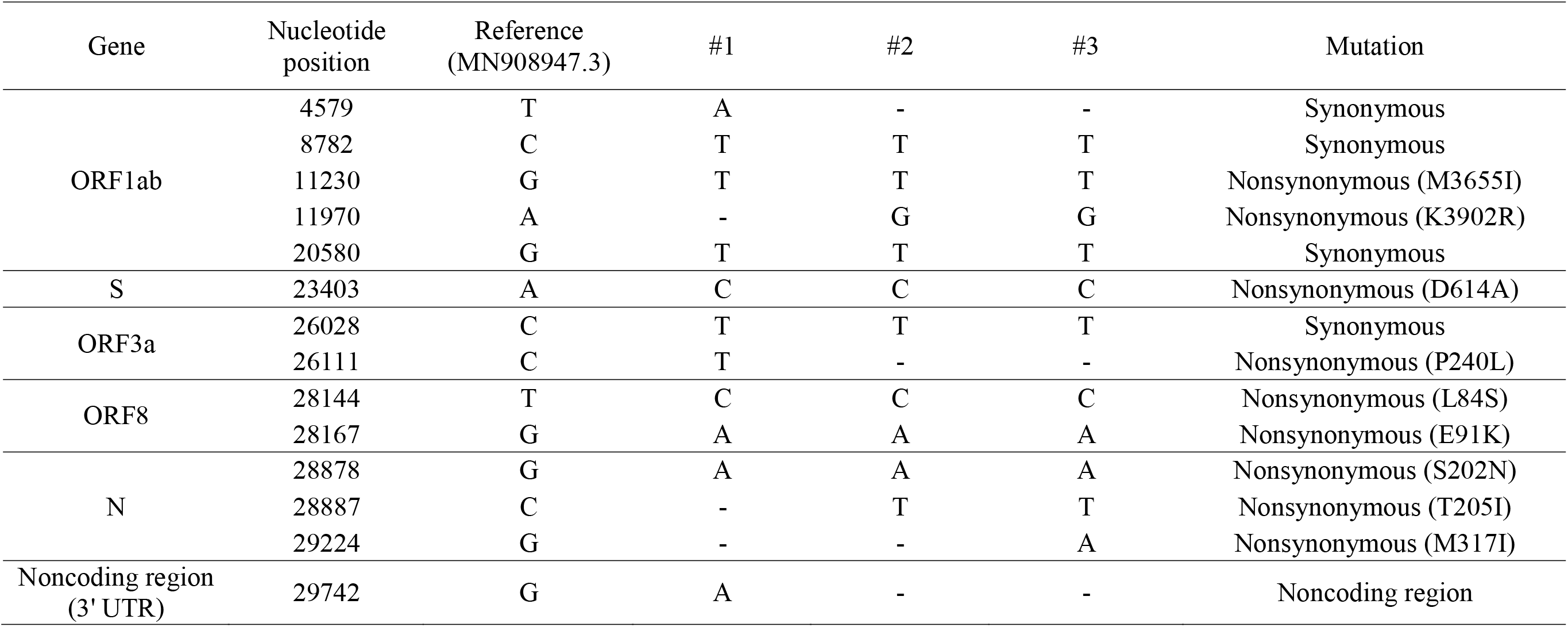
Single-nucleotide variants (SNVs) of SARS-CoV-2 isolates from travelers returning to Korea from Uzbekistan with respect to the reference genome

In addition, two travelers who visited Uzbekistan and returned to Korea were diagnosed with COVID-19 immediately after entering Korea. They arrived in the South Korea on July after short-term travel (Supplementary Table 1). The WGS results of the above two isolates also indicated that they had D614A mutations (Table 1). Due to the lack of WGS data on isolates from Uzbekistan in GISAID, we were unable to compare our mutation results to those from Uzbekistan. However, the identification of D614A in the S protein from three individuals who traveled Uzbekistan in recent days emphasized that new types of mutations might be emerging and spreading in that area.

The significance of the D614G mutation of the S protein for SARS-CoV-2 infection was elucidated in a recent report by Korver *et al*. (5). Korver *et al*. suggested that an association exists between the G614 variant and higher levels of viral nucleic acid in the upper respiratory tract in patients, which make it more infectious than D614 and has allowed the rapid spread of G614 throughout the world. Additionally, this higher level of infection might require a higher level of antibody, which is correlated with vaccines or antibody-based therapeutic agents. From a structural perspective, the D614G mutation destabilizes the interaction between the S1 and S2 units, resulting in the promotion of S1 shedding. Additionally, this mutation induces the conformational change of the receptor-binding domain (RBD) of the S protein allosterically to promote the binding of angiotensin-converting enzyme 2 (ACE2) (6). Additionally, the D614G mutation may modify the secondary structure of the furin cleavage site to facilitate the cleavage of S1 and S2 (7). Considering that the D614G mutation is expected to have significant effects on the spread of SARS-CoV-2, it will be necessary to study and characterize the D614A mutation to evaluate its phenotypes.

In this study, we identified novel D614A mutations in SARS-CoV-2 isolated from travelers returning to Korea from Uzbekistan in recent days. The results of this study emphasized the need for enhanced surveillance, which will be essential to combat COVID-19, especially where genetic epidemiology data are scarce.

Dr. Park is a senior researcher at the Korea Centers for Diseases Control & Prevention. Her research interests include the molecular epidemiology of viral and bacterial diseases.

## Data Availability

The source data for the Table along with the Supplementary Table presented in this paper are available upon request.

## Notes

### Competing Interest Statement

The authors have declared no competing interest.

### Clinical Trial

This study did not require clinical trials.

### Funding Statement

This work was supported by a grant from the Korea Centers for Disease Control and Prevention [4837-301-210]

